# Optimal reactive balance training characteristics post-stroke: secondary analysis of a randomized controlled trial

**DOI:** 10.1101/2025.07.24.25332137

**Authors:** Júlia O. Faria, Cynthia J. Danells, Elizabeth L. Inness, Avril Mansfield

**Author notes:** **Corresponding author:** Avril Mansfield, Toronto Rehabilitation Institute, University Health Network, Toronto, Ontario, Canada; 550 University Ave, Toronto, ON M5G 2A2.

## Abstract

**Background and purpose:** Reactive balance training (RBT) has shown promise for enhancing reactive balance control and reducing falls post-stroke. However, the optimal training parameters (e.g., intensity, duration) are unknown. This study aimed to investigate the relationship between different reactive balance training characteristics and improvements in reactive balance control and fall rates.

**Methods:** People with chronic stroke completed up to 12 one-hour reactive balance training sessions, twice per week. Training included experiencing losses of balance due to internal or external perturbations while performing voluntary tasks. The tasks were of four types: stable, quasi-mobile, mobile, and unpredictable, each with choice of three difficulty levels (normal, increased, or reduced). We analyzed the relationships between training characteristics (total number of perturbations, difficulty levels, perceived level challenge, and success rate) and fall rates post-training and changes in the reactive balance control sub-score of the mini-Balance Evaluation Systems Test (mini-BESTest).

**Results:** A higher number of perturbations was significantly associated with better post-intervention reactive balance scores on the mini-BEST (p=0.010). There were no significant associations with any other training characteristics and post-intervention mini-BEST Scores. For falls in daily life there was no significant association between any training characteristic.

**Discussion:** Greater exposure to RBT was associated with improvements in reactive balance control among individuals with chronic stroke. Participants who completed more sessions, and consequently experienced more perturbations, achieved better outcomes. These findings highlight the importance of sufficient training volume, suggesting that a higher number of perturbations may be optimizing the effects of RBT in stroke rehabilitation.

**Trial registration:** ISRCTN05434601

## INTRODUCTION

Stroke leads to a range of functional motor impairments that negatively impact quality of life (Poomalai et al., 2023), and is one of the leading causes of adult disability worldwide (Donkor, 2018; Saini et al., 2021). Recovering balance and independent walking form the cornerstone of improving quality of life post-stroke, and are consequently a primary focus of stroke rehabilitation (Kinoshita et al., 2017; Zhang et al., 2022).Preventing a fall after a loss of balance requires the body to respond quickly and effectively with reactive strategies (Maki & McIlroy, 1997). However, these responses are often impaired after a stroke; people with stroke may exhibit multi-step responses, delays in step initiation, and/or incomplete stepping (Gray et al., 2019; Handelzalts et al., 2024; Inness et al., 2014; Lakhani et al., 2011). Currently, many interventions aimed at improving balance post-stroke focus predominantly on anticipatory strategies (Ahmed et al., 2021; Hyun et al., 2021; Rose et al., 2017).

While beneficial to improving balance, these approaches likely do not translate to the ability to react to unexpected balance perturbations (Mansfield et al., 2018; Wang et al., 2023). Reactive balance training (RBT) employs balance perturbations to trigger rapid reactions to regain stability in a safe and controlled environment, with the goal of improving reactive balance control (Devasahayam et al., 2022). Results of clinical trials to date are promising, supporting the use of RBT post-stroke. After six weeks, reductions in the number of extra steps (Schinkel-Ivy et al., 2019) and improvements in the reactive section of the mini-Balance Evaluation Systems Test (mini-BEST; Mansfield et al., 2018) were observed after RBT compared to a control group. Dusane & Bhatt (2022) found that exposure to just 10 slips—eight on the non-paretic side and two on the paretic side—was sufficient to reduce fall incidence in the laboratory. Despite the limited dose of training, participants improved effectiveness of balance reactions, reduced falls, increased reactive step length, and had more favourable slip kinematics on the non-paretic side. However, these previous studies also raise questions regarding the optimal dose and delivery of RBT. Several studies have highlighted substantial variability in training characteristics, including number and type of perturbations, frequency, intensity, and duration (Devasahayam et al., 2022). In clinical practice, this variability contributes to uncertainty among clinicians about how to implement RBT effectively and safely. For instance, Jagroop et al. (2022) reported that many rehabilitation professionals are unsure about how many perturbations are needed to induce meaningful improvements, or how to tailor the training to individual capabilities. Understanding these aspects is crucial for designing more personalized and effective interventions. In this context, the present study aimed to examine the relationship between the characteristics of RBT and improvements in reactive balance control and fall rates in individuals with chronic stroke.

## METHODS

### Study Design

This study involved secondary analysis of a multisite randomized controlled trial (RCT) (Mansfield et al., 2018; Mansfield et al., 2015) clinical trial registration number: ISRCTN05434601). The RCT was approved by the University Health Network Research Ethics Board.

### Participants

Participants were eligible for inclusion in the original study if they were able to stand independently without upper limb support for more than 30 seconds and tolerate at least 10 balance perturbations. Exclusion criteria were: height > 2.1 m and/or weight > 150 kg; other neurological conditions; lower limb amputation; inability to comprehend instructions in English; significant illness, injury, or surgery within the past six months; severe osteoporosis; poorly controlled diabetes or hypertension; participation in physical therapy or supervised exercise programs targeting balance and mobility between recruitment and post-training assessment; and/or prior participation in RBT within one year of enrollment. Participants were included in the current analysis if they were assigned to the RBT group at the Toronto Rehabilitation Institute site; completed both the initial and final assessments, including the mini-BEST, Berg Balance Scale; and reported falls in daily life after the intervention. The researcher responsible for the pre- and post-training assessments was blinded to group allocation. Participants provided written informed consent prior to participation in the RCT.

### Intervention

The intervention consisted of two-1 hour sessions per week for 6 weeks. Sessions were delivered on a one-on-one basis (one physiotherapist per participant) in a research laboratory setting. The laboratory was equipped with a 2.63 × 2.63 m four-post XY patient lift gantry (Prism Medical, Concord, Ontario, Canada). RBT sessions included a 5-to 10-minute warm-up, voluntary tasks designed to induce internal balance perturbations, voluntary tasks combined with external balance perturbations, and a cool-down period. For safety during the exercises, participants wore a custom safety harness (ABG Concept Medical, Valcourt, Quebec, Canada) attached to the overhead support system. Internal perturbations occurred when participants failed to maintain balance during a voluntary task, such as kicking a soccer ball. External perturbations were caused by external forces, such as being pushed by the physiotherapist. The 12 training sessions were distributed such that balance losses were experienced during the following voluntary task types: 2 sessions of stable tasks like lean-and-release or body sway over stable base of support; 4 sessions of quasi-mobile tasks like stepping forward with alternate feet or walking on the spot; 4 sessions of mobile tasks like rapid turning on the spot in cued direction, or walking from one location to another; and, 2 sessions of unpredictable tasks like four square stepping to unpredictable physiotherapist-cued square). The goal was for participants to experience at least 60 balance perturbations per session.

The intervention followed a general guideline and was individualized to match each participant’s abilities. Tasks could be modified to increase or reduce difficulty as needed. Physiotherapists documented activities performed in each session, the perceived level of challenge, adverse events, any deviations from prescribed activities, adaptations to increase (e.g., closing eyes, reducing the size of the base of support, adding an unstable surface and/or increasing the height of the obstacles) or decrease (e.g., increasing the size of the base of support, taking short steps, using a large ball for throwing, and/or slowing down speed) the difficulty of the task, and participant response to the perturbations. A successful response to a perturbation was defined as requiring ≤2 steps to successfully recover balance, while failure was characterized by multiple steps, a ‘fall’ into the safety harness, or needing assistance from the physiotherapist to prevent a fall. The complete training protocol is available elsewhere (Mansfield et al., 2018).

### Data analysis

Multiple linear regression (Section II of the mini-BEST) and negative binomial regression (fall rate) were used to determine the relationship between training variables and improvements in reactive balance control and falls in daily life, respectively. There were six models for each dependent variable, with independent variables: 1) total number of perturbations completed; 2) number of perturbations completed at each training difficulty level (reduced, normal, or increased difficulty); 3) percentage of total perturbations for each type of task completed (Stable Tasks, Semi-mobile Tasks, Mobile Tasks, and Unpredictable Tasks); 4) mean perturbation success rate; and 5) mean perceived level of challenge. Pre-training mini-BEST Section II score was included as a covariate in all linear regression models. Follow-up time was included as an offset variable in the negative binomial regression models. Total number of perturbations, in units of 100 perturbations, was also included as a covariate in models 2-5. Statistical analyses were performed using Stata (Stata/SE 18.5, StataCorp LLC). Results were reported as estimated coefficients, 95% confidence intervals, and statistical significance set at p < 0.05.

## RESULTS

### Participants

Initially, 41 participants were allocated to the RBT group. However, 11 participants were excluded from the analysis: two did not complete the post-training assessment, and nine were recruited at another site. The final sample of participants is presented in Table 1.

**Table 1.**
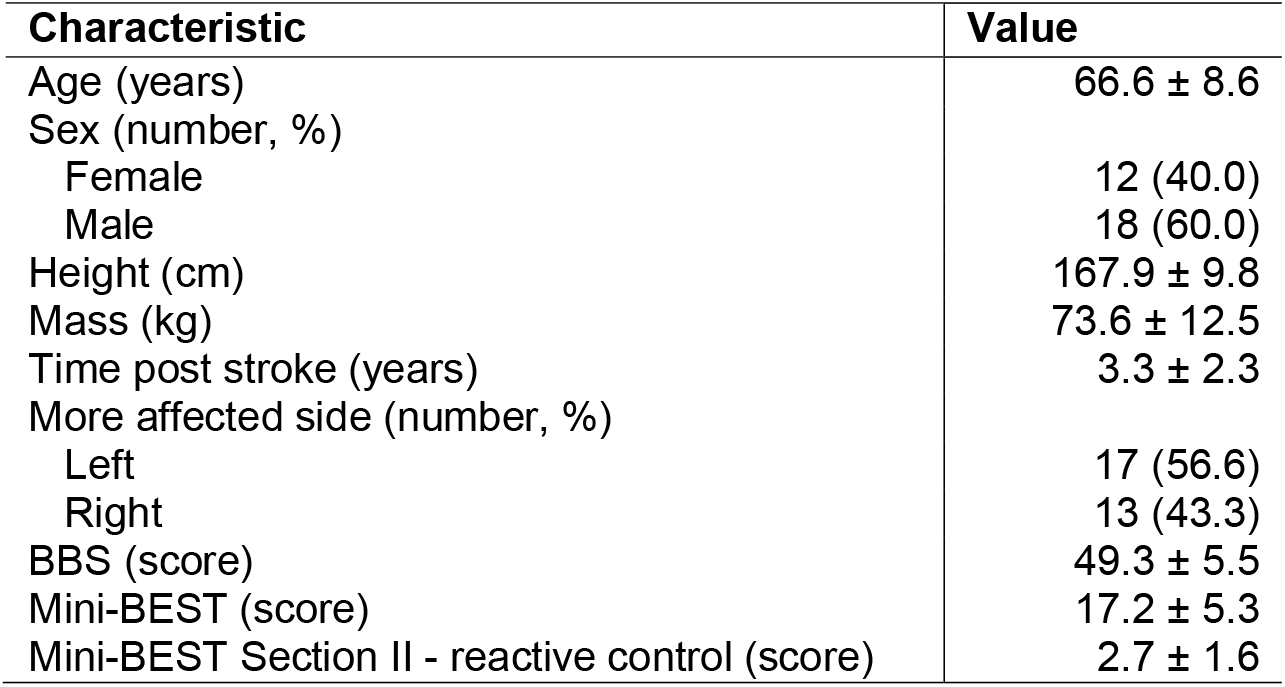
Baseline participant characteristics. Values are expressed as mean ± standard deviation for continuous variables or count with the percentage in parentheses for categorical variables.

#### Reactive balance

A higher number of perturbations was significantly associated with better post-intervention reactive balance control scores in the reactive balance control sub-score of the mini-Balance Evaluation Systems Test (mini-BESTest); for each additional 100 perturbations completed during training, the mini-BEST score increased by an average of 0.48 points (p=0.010). There were no significant associations with any other training characteristics and post-intervention mini-BEST Scores (Table 2).

**Table 2.** Multiple linear regression results for reactive balance control (Section II of the mini-BEST). Estimates are slopes, controlling for mini-BEST Section II scores at baseline and total number of perturbations (all models except total number of perturbations), with 95% confidence intervals in brackets.

|  | Estimate | p-value |
| --- | --- | --- |
| Model 1: Number of perturbations (/100 perturbations) | 0.48 [0.08, 0.89] | 0.010* |
| Model 2: Task difficulty |  |  |
| Number of perturbations with reduced difficulty (/100 perturbations) | -0.57 [-1.58, 0.42] | 0.24 |
| Number of perturbations with 'normal' difficulty (/100 perturbations) | -0.11 [-1.12, 0.90] | 0.81 |
| Number of perturbations with increased difficulty (/100 perturbations) | 0.15 [-0.48, 0.80] | 0.61 |
| Model 3: Type of task |  |  |
| Stable tasks (% trials) | 0.24 [-0.12, 0.60] | 0.18 |
| Semi mobile tasks (% trials) | 0.20 [-0.15, 0.56] | 0.25 |
| Mobile tasks (% trials) | 0.09 [-0.24, 0.44] | 0.56 |
| Unpredictable tasks (% trials) | 0.08 [-0.27, 0.43] | 0.63 |
| Model 4: Perceived level of challenge | -0.39 [-1.26, 0.47] | 0.36 |
| Model 5: Success rate (% of trials) | 0.00 [-0.03, 0.03] | 0.99 |

#### Falls in daily life

There was no significant association between any training characteristic and falls in daily life (Table 3).

**Table 3.** Negative binomial regression results for fall rates. Estimates are incidence rate ratios, controlling total number of perturbations (all models except total number of perturbations), with 95% confidence intervals in brackets.

|  | Estimate | p-value |
| --- | --- | --- |
| Model 1: Total of number of perturbations (/100 perturbations) | 1.10 [0.73, 1.65] | 0.63 |
| Model 2: Task difficulty |  |  |
| Number of perturbations with reduced difficulty (/100 perturbations) | 1.23 [0.43, 3.52] | 0.69 |
| Number of perturbations with 'normal' difficulty (/100 perturbations) | 1.52 [0.68, 3.66] | 0.28 |
| Number of perturbations with increased difficulty (/100 perturbations) | 0.90 [0.42, 1.93] | 0.79 |
| Model 3: Type of task |  |  |
| Stable tasks (% trials) | 0.90 [0.62, 1.13] | 0.61 |
| Semi mobile tasks (% trials) | 0.97 [0.67, 1.41] | 0.89 |
| Mobile tasks (% trials) | 0.87 [0.62, 1.23] | 0.45 |
| Unpredictable tasks (% trials) | 1.06 [0.72, 1.56] | 0.74 |
| Model 4: Perceived level of challenge | 0.48 [0.19, 1.17] | 0.10 |
| Model 5: Success rate (% of trials) | 1.01 [0.98, 1.05] | 0.33 |

## DISCUSSION

This study aimed to determine the relationship between the characteristics of RBT and improvements in reactive balance, and falls during activities of daily living in individuals with chronic stroke. While RBT has shown promise in reducing falls and enhancing reactive balance control, which are key targets for this training modality (Handelzalts et al., 2019; Mansfield et al., 2018, 2018; Rieger et al., 2024), there is still a gap regarding the optimal dosage. Specifically, little is known about how the number of perturbations or the total time of training exposure influences outcomes, which limits the development of clear clinical recommendations (Hezel et al., 2025). This study addresses that gap by exploring how different levels of exposure to perturbations relate to improvements in reactive balance control.

Participants who experienced more perturbations achieved greater improvements in a measure of reactive balance control. Classical models of motor learning support that, while the greatest gains often occur during early stages of practice, continued improvement requires prolonged exposure until the task becomes automated (Crossman, 1959; Newell, & Rosenbloom, 1981). These findings have been applied to stroke rehabilitation, where it is recommended that thousands of repetitions of a motor task are required to bring about lasting changes in skill performance (French et al., 2016; Vearrier et al., 2005). Indeed, studies have reported that a greater number of repetitions is often required to achieve functional improvements post-stroke compared to healthy individuals (Cirstea et al., 2003; Winterbottom & Nilsen, 2024; Sousa et al., 2019). This may help to explain the association observed in our study between greater exposure to perturbations and improved reactive performance, supported by neurophysiological mechanisms involved in motor adaptation (De Sousa et al., 2019; Hao et al., 2022; Levin & Demers, 2021). Repeated exposure to balance perturbations allows the central nervous system to refine feedback-based reactive responses and incorporate feedforward control strategies, leading to more efficient sensorimotor processing (Pai & Bhatt, 2007). Supporting the notion that a sufficient number of perturbations is necessary to elicit meaningful improvements, a previous study using 12 training sessions of 66 perturbations per session in post stroke participants—comparable to the dose used in our study—reported significant improvements in multiple-step thresholds and balance confidence (Handelzalts et al., 2019).

In contrast to our results, others have found lasting improvements in reactive balance control with a very low volume of RBT among healthy older adults (e.g., a single session of 24 perturbations; (Bhatt et al., 2012; Pai et al., 2014). The authors suggested that learning could be reinforced by perceived or actual penalties resulting from recovery failures, thereby motivating the central nervous system to learn quickly (Sacchetti et al., 2004), particularly in people with a previous history of falls. Studies have also demonstrated improvements in reactive balance control post-stroke with low training doses (e.g., one session of 24 perturbations Bhatt et al., 2012; Chayasit et al., 2022). The findings of these previous studies suggest that large effects may be observed with a very low volume of RBT. However, using correlation analysis, our findings suggest that people with stroke can achieve more benefits with increased volume of RBT. No studies have directly compared the effects of different volumes of RBT; therefore, our results will need to be confirmed with randomized controlled trials.

We also explored whether variations in training challenge were associated with improvements in reactive balance control and reductions in daily-life falls. Specifically, we examined both the absolute challenge level (i.e., adjustments to task difficulty and progression from stable to unpredictable tasks) and the relative challenge level (i.e., ‘success rate’ and perceived difficulty).

Although these elements may represent aspects of training “intensity”, we acknowledge that intensity of balance training is not straightforward to define (Farlie et al., 2013). In this study, we operationalized intensity as the proportion of trials eliciting multi-step responses, targeting approximately 50% across participants. This approach may have limited variability in relative intensity between participants. In contrast to recommendations that higher-intensity balance training yields greater reductions in fall risk (Sherrington et al., 2017), we found no association between these indicators of training challenge and the observed outcomes.

Training in this study was highly variable in terms of perturbation types (internal and external), directions, timing, and concurrent tasks. Although this variability was not systematically controlled, it may have contributed positively to learning outcomes. According to motor learning theory, introducing variability during practice—particularly in the acquisition phase—can enhance long-term retention and transfer of skills, even when initial performance is reduced (Hanlon, 1996; Reisman et al., 2007).

Variable practice has consistently demonstrated superior outcomes compared to constant or blocked paradigms, both in neurologically intact individuals and in stroke populations (Reisman et al., 2009; Tyrell et al., 2014). In neurorehabilitation, this type of training is associated with improved generalization and adaptability, which are key goals when working with individuals who must relearn balance strategies (Shea & Morgan, 1979). Such variability is also consistent with established motor learning principles, which suggest that it promotes adaptability and more robust motor representations (Schmidt & Lee, 2011; Shumway-Cook & Woollacott, 2017). These principles may help to explain our findings. Participants who experienced a higher number of perturbations, thus exposed to a greater variety of stimuli, achieved greater improvements in reactive balance. This suggests that variability in exposure, when combined with sufficient task repetition, may reinforce motor representations and facilitate more efficient sensorimotor responses. While feedback control remains essential in reactive balance, repeated exposure likely promotes faster and more effective responses by refining internal models (Pai & Bhatt, 2007). Our results align with this view, as improvements occurred even in the absence of a strict progression in intensity, possibly due to the diversity and volume of exposure during training. This may partially explain the improvements observed in our participants, despite the lack of a direct association with training intensity or dosage (Krakauer, 2006).

The relationship between post-training falls and specific components of the intervention was also examined, but no significant differences were found. Falls are multifactorial, influenced not only by impaired balance, but also by other individual-level factors, as well as task and environmental factors (Campbell & Matthews, 2010; Shumway-Cook, A & Woollacott, M.H., 2017; Sullivan et al., 2023). For example, increased physical activity as people improve balance and confidence in independent mobility may actually contribute to an increased incidence of falls (Aihara et al., 2021; Suzuki et al., 2005).

Future research should further explore how different task types interact with individual patient characteristics to maximize the benefits of RBT, ensuring they are both effective and tailored. Additionally, efforts should focus on identifying the optimal dosage and controlled intensity required to enhance reactive balance training in post-stroke patients. Establishing clear parameters would facilitate the distinction between different practice structures and support the replicability of protocols in both clinical and laboratory settings, ultimately contributing to the development of more effective and individualized fall prevention strategies for post-stroke patients.

### Limitations

The correlational design employed does not permit a comprehensive assessment of the protocol’s efficacy over time. Only baseline and post-training assessments were conducted, with no intermediate evaluations during the training sessions, thereby limiting our ability to characterize the learning trajectory throughout the protocol. Furthermore, while it is logical to assume that completing more perturbations during training contributed to greater improvements in reactive balance control, we cannot assume a causal relationship. For example, we cannot rule out the possibility of unmeasured variables contributing to both reduced volume of training and reduced adaptations to training.

### Conclusions

A structured RBT protocol enhances reactive balance in individuals with chronic stroke. Exposure to a higher number of perturbations—typically achieved by participants who completed more sessions— was associated with greater improvements in reactive balance control, suggesting that increased training volume may be critical for optimizing outcomes. While task difficulty and perceived challenge did not significantly predict improvements, our findings highlight the importance of training dosage and exposure in shaping clinical responses to RBT in this population.

### Implications for physiotherapy practice

- A higher number of perturbations was associated with better balance recovery, reinforcing the importance of sufficient exposure to reactive challenges.
- This finding provides valuable insight for professionals, supporting a higher volume of reactive balance control for better outcomes in clinical practice.
- Repeated exposure to perturbations led to meaningful improvements, suggesting that consistency and volume may be more influential than intensity alone in stroke rehabilitation.
- A protocol with varied task types and perturbation contexts may have supported motor learning and reactive control, even without systematic intensity progression.

## Data Availability

Participants did not provide consent to share data externally.

## Acknowledgments

We acknowledge Anthony Aqui for assistance with data collection and processing and Svetlana Knorr for assistance with delivering the intervention.

